# The use of a participatory patient engagement research project to meaningfully engage those with lived experience of diabetes and homelessness

**DOI:** 10.1101/2021.02.26.21252531

**Authors:** David J.T. Campbell, Rachel B. Campbell, Anna DiGiandomenico, Matthew Larsen, Marleane A. Davidson, Kerry A. McBrien, Gillian L. Booth, Stephen W. Hwang

## Abstract

**Introduction:** Participatory research is a study method that engages patient partners in research programs from study design through to completion. It has seldom been used in diabetes health services research. Our objectives were to describe the process and challenges of conducting a patient-engagement project and to highlight the experiences of patient participants and academic researchers.

**Research Design & Methods:** We recruited PWLEH and diabetes in Toronto, Canada to be patient partners. Group members were asked to commit to attending biweekly meetings. We undertook two major research projects: Concept mapping to choose a research focus; and photovoice to explore accessing healthy food while homeless. We used a convergent mixed methods design to evaluate their experience.

**Results:** A diverse group of 8 PWLEH had an average attendance of 82% over 21 meetings – despite this success, we encountered a number of challenges to conducting this research. Group members reported that participation improved their ability to be self-advocates in their diabetes care and provided them with tangible skills and social benefits. Group members stated that they valued being involved in all aspects of the research, in particular knowledge translation activities, including advocating for nutritious food at shelters; presenting to stakeholders; and meeting with policy makers.

**Conclusions:** The use of participatory patient engagement research methods enables academic researchers to support community members in pursuing research that is pertinent to them and which has a positive impact. In our study, group members contributed in meaningful ways and also valued the experience.

**What is already known about this subject?:**

- Patient oriented research is important to public health research as it helps with the development of relevant interventions and knowledge translation.
- Participatory research is a form of research that maximally involves patients in all phases of the research.
- Participatory research has rarely been used in research on diabetes and diabetes-related interventions.

**What are the new findings?:**

- Patient engagement is important for studies involving socially disadvantaged populations with diabetes.
- Community members involved in research contribute substantially to research projects but also find the experience to be enriching and valuable.

**How might these results change the focus of research or clinical practice?:**

- Those who conduct research with and develop programs to provide diabetes care, especially to socially disadvantaged populations, should involve community members through all phases of the process to ensure the intervention is maximally useful for patients.

## Introduction

The importance of engaging patients in all aspects of developing health care policies has been widely acknowledged ^1^. The definition of patient-oriented research, or patient engagement research, is broad and includes research that involves patients in a variety of capacities ^2 3^. Arnstein originally described citizen participation as a spectrum on a “ladder” of participation, ranging from simply collecting data from informants or subjects to processes where citizens are fully engaged at all stages ^4^. Typically, participatory research studies are those towards the top of this ladder – those which give patients or citizens increased levels of power in the process, including: Partnership, Delegated Power, and Citizen Control ^4^.

One particular form of patient engagement research is Community-Based Participatory Research (CBPR); wherein patients or community members are viewed as co-researchers, rather than as study subjects or participants ^5^. CBPR has been used extensively in social work research ^6^, in international settings ^7^, and more recently in health services research ^8^. While participatory research approaches have been used in the field of diabetes ^9 10^, a recent realist review found a total of only 29 studies in which patients or community members actively contributed to research projects on diabetes prevention and outcomes in more involved ways than being traditional informants ^11^.

CBPR and patient-engagement research may have a particularly important role in areas of health research focused on equity and the social determinants of health ^12^. We were interested in inequities in diabetes management and outcomes among people with lived experience of homelessness (PWLEH). PWLEH are known to have more difficulties managing their diabetes ^13^ and are more likely to accrue diabetes-related complications ^14^. There are many potential explanations for these disparities in diabetes outcomes, as PWLEH face a number of barriers including: mistrust of the healthcare system ^15^, lack of health insurance coverage ^16^, and difficulty accessing healthy foods ^17^. Furthermore, typical diabetes care models may not service this population optimally ^18^. While several studies have highlighted the difficulty of managing diabetes while experiencing homelessness, few, if any, have attempted to elicit the voice and preferences of this population with regards to these challenges.

We used participatory methods (at the Partnership level of the participation ladder) to engage PWLE of homelessness and diabetes with the goal of empowering them to lead and undertake meaningful research initiatives. The objective of this paper is to describe the process of conducting this participatory patient-engagement project and to report the experiences of individuals with lived experience of homelessness and diabetes, giving voice to patients. Our specific aims were to:

1. Document our ability to engage this population in group research activities, and the challenges inherent in conducting patient engagement research with this population.
2. Qualitative explore the experiences of group members who participated in the process.
3. Documenting reflections of group members and academic researchers on aspects of the work that were most meaningful to them.

## Methods

### Study Design

From the outset, we envisioned an open-ended participatory patient-engagement project. The patient engagement research approach taken in this work is grounded in the theoretical idea of ‘giving voice’ through social representation theory ^19^, which theorizes that “knowledge is the outcome of social interaction” ^20^. The objective of giving voice has been described as “empowering people to be heard who might otherwise remain silent” ^21^.

In order to reach our objective, we recruited a group of PWLEH and diabetes to form the Clients with Diabetes Action Committee (CDAC) and used participatory research principles to guide them in developing research priorities and pursuing meaningful research activities ^5^. We were careful to keep the research topic and question as open-ended as possible to allow research priorities and questions to emerge from within the group rather than being imposed upon group members by researchers and/or funding bodies. Ethics approval for this work was received from the Conjoint Health Research Ethics Board of the University of Calgary (REB# 18-1663), as well as the Research Ethics Board of St. Michael’ s Hospital, Unity Health Toronto (REB # 18-288).

### Setting & Group Composition

We conducted this program of research in Toronto, Ontario – Canada’ s largest city (2017 population: 2,930,000). Toronto is a city with high levels of ethnic and socioeconomic diversity ^22^. It is known for having very high housing costs ^23^, which contributes to it also having the greatest absolute number of people experiencing homelessness in Canada ^24^. Based on 2018 data, 8715 unique individuals experience homelessness on any given night in Toronto ^25^.

We sought to include individuals who had recent experience managing diabetes in the context of homelessness. Therefore, eligibility criteria to join the CDAC included a self-reported history of living with type 1 or type 2 diabetes while experiencing homelessness or housing instability at any point in the previous 2 years. We use the broad definition of homelessness endorsed by the Canadian Observatory on Homelessness: lacking “stable, safe, permanent, appropriate housing” ^26^. This includes individuals who are sleeping rough, emergency sheltered, provisionally accommodated, and precariously housed. We excluded anyone who had only diabetes risk factors, without a previous diagnosis of diabetes. Additional exclusion criteria included the inability to fluently converse in English and active severe mental illness that would preclude one’ s ability to participate in group research activities. Recruitment began in November 2018 and was complete by the end of January 2019. Participants had to commit to joining the group and attending approximately every other week from January 2019 to July 2019.

We recruited participants primarily through recruitment flyers that were posted in numerous downtown Toronto shelters, drop-ins, rooming houses, addiction recovery facilities, and community notice boards, as well as community health centres and programs, endocrinology clinics and a community-based education and addiction recovery service. The flyers clearly outlined the eligibility criteria (diabetes and homelessness) and stated that participants would be compensated. Interested parties were encouraged to either contact the investigators by telephone or email, or to attend one of the group meetings, as dates and times for several meetings were included on the recruitment advertisements.

The group met regularly in a community space in Regent Park, a lower-income area on the east side of downtown that was originally built as a public housing project in the 1940s. Participants were not necessarily residents of this area, but all were familiar with it and most lived within walking distance. Initially, the group gathered for two hours every other week. When participants attended their first group meeting, they were introduced to the purpose and structure of the group and provided written informed consent to participate. They also signed a confidentiality agreement, a Terms of Reference document, emphasizing the importance of professionalism and mutual respect towards facilitators and peers. At the first group meeting, all participants were provided a $10 gift card to a local coffee shop. To recognize the time and effort of regular group members, they were provided with 2 transit tokens and a $20 (CAD) cash honorarium for each subsequent group meeting they attended. This amount was chosen as it approximated a working wage in Ontario. We spent roughly 40 minutes of each meeting sharing a meal and providing diabetes education, so we sought to compensate people for 80 minutes. With a local minimum wage of $14CAD/hr, our $20 stipend corresponds to a working rate. Furthermore, participants were compensated at the same rate for work done outside group time.

### Group training

Meetings were facilitated by two investigators (DJTC and RBC), who were trained and experienced in qualitative data collection methods and in working with PWLE homelessness. Neither had pre-existing relationships with group members.

Initial group meetings began with a review of the Terms of Reference each member had signed. Group members and facilitators were encouraged to get to know one another using ice breakers and participating in social activities. During this part of the group meeting, coffee and a nutritious lunch was served to group members.

In order to empower group members to become fully engaged in the research effort, we utilized several of our early group sessions to provide basic training in the purpose, fundamental principles, and methods used in academic health research. We employed and modified resources that others had utilized for participatory research in other unrelated contexts ^27^. During this process, group members received introductions to study methods used in participatory research, including designing surveys, conducting interviews, and using arts-based methods.

Many group members had not received basic diabetes education in the past, and/or lacked sufficient knowledge and understanding about diabetes management and complications to contextualize the personal challenges they and other members of their community were facing. Since this is a necessary step to enable the group to reach consensus on topics fit for research, we planned that diabetes education would be a recurring core element of group meetings. The remainder of group meeting time was devoted to pursuing the scholarly activities proposed by the group (as described in detail, below) and making plans for future meetings.

### Research Activities

Having community members decide which research questions are most important to address is a fundamental component of participatory research. At the outset it was clear that group members each had priorities and interests that were primarily driven by their own experiences of managing diabetes while experiencing homelessness. In order to concisely synthesize the group’ s priorities, we used a participatory research methodology known as concept mapping, which has been used extensively to help identify priorities for particular groups ^28^. The details of the concept mapping process and results will be published separately. Through an iterative process which included brainstorming, sorting, rating, and discussion, our group members identified that the collective preference for our group’ s research activity would be on how homelessness affects one’ s ability to access healthy foods which, in turn, impacts diabetes management and outcomes.

Once the results of the concept mapping exercise were analyzed, the group facilitators presented different research methodologies that could be used to explore this topic further, including quantitative methods (such as surveys) and qualitative methods (such as focus groups), as well as arts-based methods (such as photovoice and documentary film). The group collectively decided to pursue a project based on the classic photovoice methodology, as described by Wang ^29^. The photovoice project is also documented in a separate forthcoming publication. Group members were tasked with taking photographs that illustrated each of the following specific research questions:

> *What makes it challenging to eat well with diabetes while experiencing homelessness?*

> *What about homelessness has been a major help or barrier to diabetes self-management?*

Group members then showed their photos to each other and to the facilitators. They chose which photo they wanted to use and developed a narrative to accompany the photo, aided by the facilitators using photo elicitation techniques ^30^.

The photos and narratives resulting from the photovoice project were mounted and framed. They were displayed alongside the biographies of the photographers. The photo exhibit, and descriptions of this program of research were presented at local hospitals, research institutes, and public exhibition spaces in Calgary and Toronto, in addition to national conferences including: Diabetes Canada (Winnipeg, MB, Oct 2019), Canadian Alliance to End Homelessness (Edmonton, AB, Nov 2019), North American Primary Care Research Group (Toronto, ON, Nov 2019).

### Evaluation methods

We undertook a convergent mixed methods evaluation of our group research activities after completion of the study. First, we gathered information on attendance of group members at meetings. We also sought to allow them to provide feedback on the experience and to understand the challenges inherent in conducting this type of research with this population. To do so, we conducted anonymized surveys (Appendix A) and an open-ended focus group at the conclusion of the group meetings in July 2019, with the latter facilitated by colleagues not previously engaged with the group. After a 4-month hiatus, we recalled group members and conducted a final semi-structured interview to ask them about how their participation impacted their lives, and what might have changed for them. Interview responses and focus group transcripts were analyzed using directed qualitative content analysis ^31^, following a predominantly deductive approach with the coding template being based largely upon the questions asked in the quantitative survey. Results from the various methods were triangulated to provide qualitative support for the patterns reported in the quantitative survey. Finally, we asked some group members and the academic investigators who were closely involved with data collection to provide a statement reflecting on their experience and the impact of their participation in this research.

## Results

### Group composition, attendance, and challenges

We were contacted by 28 individuals who initially expressed interest in being part of the group. Sixteen (16) of those individuals came to at least one group meeting. Of those who attended one meeting, 5 were ineligible to participate as they did not have diabetes, based on self-report. The remaining 11 individuals agreed to become part of the recurring group. Three participants did not return after the initial group meeting, and 8 continued to participate throughout the entire duration of the study. Our 8 group members included 5 women and 3 men, with ages ranging from 36 to 73 years. Four reported having diabetes complications while 4 stated that they had no complications. The majority of group members were white (n=5), two were black and one was Indigenous (Table 1).

**Table 1:**
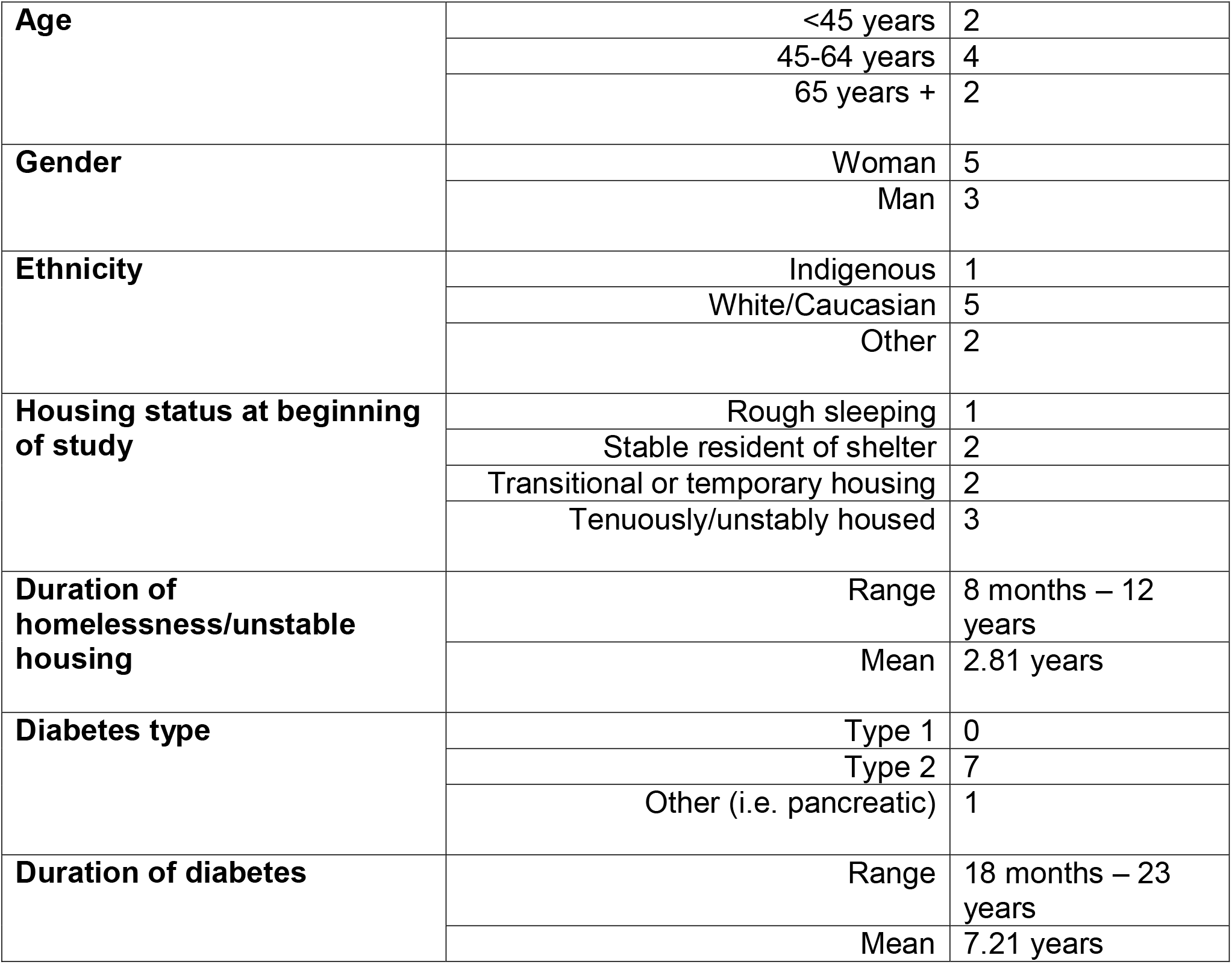
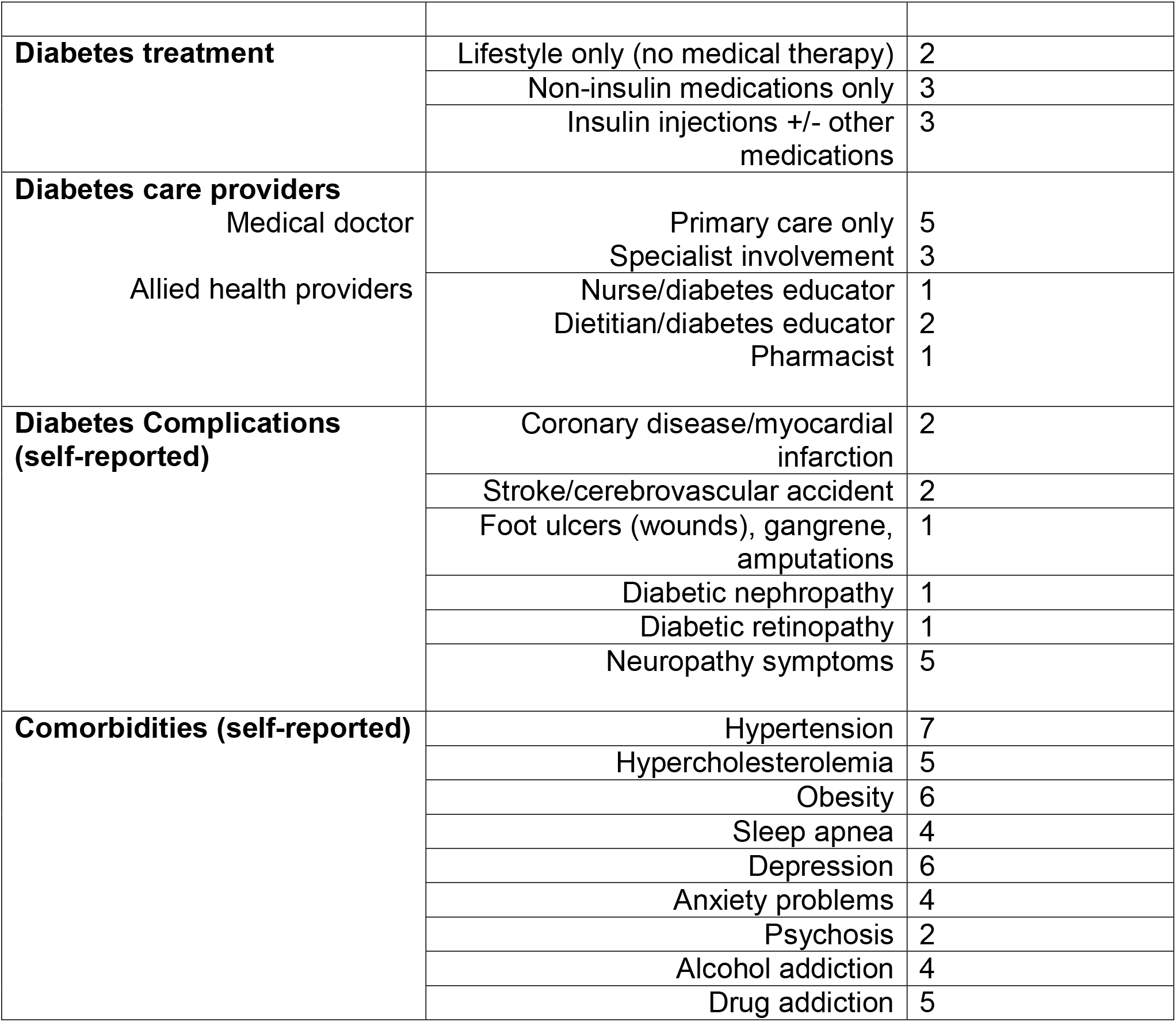
Group Member Characteristics.

The attendance rate for the 8 regular group members was 82%. Individual attendance ranged from 48% (10/21 meetings) to 100% (21/21 meetings), with an individual mean of 80%. Two academic investigators (DJTC and RBC) attended all group meetings, teaching the diabetes education portion of meetings, and facilitating group discussions and activities during the remainder of the group time.

We noted a number of challenges that occurred during the conduct of this research. When some regularly attending group members missed meetings we later discovered they had been in hospital. Half of our group members had health issues that caused them to visit emergency departments during the 6 months of the group, with 3 of them having three or more acute care visits in that time. Another challenge in working with this population was difficulty maintaining contact with group members. Four out of the eight group members lost or had their cellphones disconnected during the study period. Similarly, 5/8 group members moved residences during the 6-month period, some multiple times. Future studies could consider providing participants prepaid cell phones or minutes to facilitate ongoing contact.

### Participant Experience

Our evaluation activities, including surveys and question guides, focused around three areas of interest to help understand participants’ experiences of the program. These included: (1) improvements in diabetes knowledge and self-efficacy; (2) acquisition of tangible skills and benefits from participation; and (3) challenges and suggestions for improving the group research experience. The convergent results from both quantitative and qualitative data are presented together below.

#### Diabetes knowledge & self-efficacy

Seven out of eight group members rated their knowledge of diabetes as poor (4/8) or fair (3/8) prior to participating in the group. One group member stated about his prior diabetes knowledge: *“it was very poor, I didn’ t know much. Just that basically high blood sugar and low blood sugar, all of them are bad”* (P1). Another stated: *“I didn’ t know anything at all about diabetes – nor was I interested in learning anything about it”* (P7).

After the group, all of these individuals rated their knowledge as good (1/8), very good (5/8) or excellent (1/8). Participants described having acquired knowledge about diabetes complications: *“it’ s not just the sugar, but it’ s also foot care and your eyes, and overall nerve damage… before I wasn’ t aware of all that stuff”* (P5); and the pathophysiology of how these problems arise: *“damage to the blood vessels like the scarring, can lead to a build up of plaque and all kinds of problems”* (P1). Several participants remarked that they gained an appreciation of the importance of diabetes self-management: *“the more you do to address the complications that diabetes can cause, I think the more healthier it will make you”* (P2).

Furthermore, group members expressed a variety of changes that happened with their diabetes care and self-management as a result of things they learned during the group’s diabetes education sessions. One said that they were now: *“Connecting with specialists and doing the maintenance”* (P4), while another said that they had: *“added [new medication], joined a weight loss clinic, and taking fasting blood sugar measurements more often”* (P1). Another group member stated: *“I think I will be looking into joining diabetes education centres”* (P2), as a result of their participation in the group sessions.

Finally, a number of group members felt that their participation in group activities increased their ability to self-advocate for their healthcare needs. One stated: *“I would say it gave all of the different pieces of information. It helped me establish my priorities for me to pursue with my doctor”* (P2). Another said:

> *[the group] taught me how to ask what I want. I realized I’ m in control of my own healthcare and I have to ask specific questions, right? I have to be the one that has to be up there jumping up and down and asking for what I want or what I need… to actually have a doctor here that we can pick their brain you know, it was actually beneficial*. (P4)

#### Tangible skills and social benefits to participants

All group members identified that they gained tangible skills, which included: working together with a group, learning photography skills, and contributing to academic research projects. However, the most valued aspects of participation were the social opportunities afforded by the group. A group member stated that: *“Camaraderie among group members was great”* (P1). Others formed social connections that persisted outside of the group: “*any time I think that I might be eating something that I shouldn’ t be eating, I call one of the group members and be like – yo can I eat this?”* (P4).

Finally, one said: *“I was in crisis, you know, and was not getting the help that I needed from the agencies, but I think coming to this group kept me going, you know, because I was doing something and I was learning”* (P6). Finally, one summed up the social impact of the group by stating: *“we are not alone and we supported each other and we laughed, we giggled, we cried… we became friends”* (P5).

One individual described how participation in the group helped them feel like they were contributing meaningfully: *“it shows that I can do something like to help the homelessness and the diabetics. Like I can be of some help to society, generally some help instead of just sleeping all day in the shelters*” (P7). Another stated: *“I learned that for me it’ s about giving, sharing… giving my story out there, right… being there to help others, you know”* (P4).

#### Challenges and suggestions for improvements

As mentioned above, a number of group members identified that camaraderie was a strength of the group, however, the confluence of personalities also posed a challenge at times. One participant stated: *“I hated sometimes being stuck in a room for two hours with people I didn’ t get along with”* (P4). To deal with these challenges, some group members suggested having more clear expectations: *“so the boundaries are pretty clear like what we are talking about, just so people don’ t get super upset or like everybody feels like they have a voice”* (P4), or reiterating these expectations more clearly at the outset of each group meeting. Another group member suggested that group interactions may have been aided if facilitators had formal training in trauma-informed care or social work practice.

One participant stated that participating in the group was somewhat triggering and would have been strengthened by having more resources on hand: *“when I was doing the photo it dragged up a lot stuff that I was remembering being homeless and going through stuff and I don’ t have quite the support that I should have”* (P6).

Similarly, another participant mentioned that participating in the photovoice exhibit was difficult: *“I was overwhelmed, excited, just overwhelmed. It was very nervous for me, but turned out good in the end”* (P3).

Group members commented that while they appreciated the food provided during meetings they would have valued more diversity and that it was a missed opportunity for dietary education: *“when it comes to the basic foods that we actually ate we never actually looked at the array of foods that diabetics can eat and how that can be, how you can actually use those kinds of foods”* (P2). Other related feedback was that cognitive behavioural therapy approaches could have been implemented to help participants with heir diabetes self-management.

### Group Member and Academic Investigator Reflections

#### DJTC

Being a diabetologist with a clinical practice focused on caring for those who face social vulnerabilities (many of whom have been homeless), I thought that I had a good sense of the types of barriers my patients face and what their priorities would be. However, doing this patient-led project taught me how absolutely critical it is to meaningfully engage those with lived experience, as their priorities are likely to be different than what a provider might anticipate them to be. The findings from this study continue to inform my ongoing research and advocacy efforts. Given the centrality of access to healthy foods raised by this research, I am creating new programs and initiatives with the aim of addressing food insecurity in patients with diabetes. Also, as the coordination of healthcare visits was raised by the group, I am now pursuing a new line of research to pilot test a comprehensive diabetes care model in those experiencing homelessness to reduce this burden for patients similar to those in our group.

#### RBC

I have been working in homelessness policy and research for 15 years. Though I have never been a front-line service provider, I have worked with clients in focus groups, advocacy groups, and during research events. My participation in this community-based research project taught me that researchers and policy makers need to work with individuals with lived experience of homelessness on an individual, face-to-face level, regardless of their role or position. The lessons I learned from CDAC members through sharing the nuances of their lived experience are indispensable. Despite reading academic literature and having a pulse on the world of homelessness, I found the work that the group did to be deeply moving– and still do every time I revisit it. The development of the group’ s research priorities and questions serves as a constant reminder that academics are not necessarily on the same page as the those with lived experience in this regard. Academics will never know what clients/patients prioritize unless they engage with the community and enable individuals to partner meaningfully in research endeavors.

#### AD

At 56 I found myself homeless and fighting diabetes. That’s when I joined an amazing group of people, the Clients with Diabetes Action Committee. I was given a voice from them. The group was made up of all walks of life but we shared one common ground, we all wanted to be involved in a cause that touched our lives, it made me feel that my story was important. We were involved in choosing the topics that were important to us and being able to share our stories with the other members of the committee. It was an empowering experience knowing that I was not alone.

Since getting involved with the CDAC, I have been able to share my experience on a podcast ^32^ and in November 2019 I was given the opportunity to present our research at the CAEH two national conferences, one on diabetes and one in homelessness. Being able to share my experience with other researchers and doctors, ignited a fire within me. There I also had the pleasure of meeting the federal politician from my local constituency and was able to share my experience with him. He was interested in listening to my story. When I returned to Toronto he followed up with me. He directed me to other politicians and before COVID he was helping me to bring our Photo Exhibit to City Hall to tell our story. This experience have me a voice that has kept me focused on fighting for my rights. I will never let the light go out.

#### ML

Information about diabetes from the CDAC group facilitators deepened my understanding of how my diet and level of activity have a direct impact on my health outcomes. In talking with other group members I learned of helpful services that I could utilize, such as food banks with access to fresh produce and drop-ins serving nutritious food. More so, listening to group members share about their personal journeys with diabetes and homelessness, I began to understand how one can exacerbate the other. In learning about different research methods and contributing photos and writing about my experiences, I came to understand that my contribution to the study was legitimate. Brainstorming as a group with my peers on a research question demonstrated to me that the facilitators were interested in addressing issues that mattered to myself and others who have had a similar journey. In November 2019 I was able to attend and present at the Canadian Alliance to End Homelessness conference. I also had the opportunity to share my experience with the CDAC in an interview with a reporter ^33^. Talking to professionals allowed me to bring attention to issues that are important to me and demonstrated that my lived experience is valuable to people who were in a position to help others like myself.

#### MAD

My experience with the CDAC group was challenging and educational. Within the classroom sharing our experiences as individuals without appropriate housing and chronic health history was quite triggering. We all had very challenging disclosures and at times there were behaviours. I really enjoyed attending the various venues outside of the classroom. When we did our photo exhibits, this was a great opportunity to have my voice heard. I met many people that were unaware of what individuals without appropriate housing are faced with or the many different reasons we all had for being on the streets as an adult or youth. The people I spoke with were unaware that youths deal with violence and abusive situations and some adults are graduates and are employed. This was my comfort level, speaking with total strangers that I would never see again, this was a lot better than engaging with my classmates that I saw weekly.

I also enjoyed when we had guest speakers; a representative from Diabetes Canada spoke to our group and was open to a Q&A after. I spoke about my own experiences coping with type 2 diabetes and I got a chance to co write an article in a national magazine,^34^ and another in a national newspaper ^35^. I really wished that I had more opportunities to connect with policy makers. I believe that policy makers have no idea the barriers they create when they implement guidelines and rules. I am sure they have the statistics but it seems that they really don’t care.

#### SWH

I have worked in homelessness-related health research for over 20 years and have led a number of projects related to diabetes. Being involved with this project, however, was a unique experience. Having individuals who have lived with homelessness and diabetes to lead the direction of the research provided new insights from a first-hand perspective and yielded meaningful results. On the heels of this patient-led program of research, my team has launched a community-based group to continue bringing the voices of people with lived experience of homelessness to research priorities at our centre. We will also use the insights from this study to help guide our homeless-focused primary care teams as they work with patients with diabetes. In this way, the efforts of the Clients with Diabetes Action Committee live on.

## Discussion

Our patient-engagement research project involved members of the community who had lived experience of homelessness and diabetes in a participatory fashion, empowering them to work alongside the academic investigators. We were able to elicit their shared priorities and subsequently explore these using photovoice. Because they were involved from the outset, group members helped translate the study findings to community decision makers. The experience was largely positive for our group members, as they described a number of benefits they received through their participation in the group, including both benefits to their health and diabetes management, as well as other tangible and social benefits. Interestingly, the benefits that were described by participants reached far beyond what was initially envisioned by study investigators at the outset of the project (i.e. enhanced diabetes knowledge, photography skills). Participants felt that some of the most important benefits they received from the study were intangibles: feeling respected, valued, and heard; having a sense of accomplishment and purpose, which led to increased self-efficacy in other domains; and developing a community of people with whom they shared many life experiences. This finding is similar to what has been seen in other studies using participatory research ^36^. Context, group dynamics, community centeredness, and research design have been elicited as key elements of participatory research ^37^. As shown in our data, each of these played a key role in the success of our project.

Patient-oriented research, or patient engagement, is becoming increasingly important in health services research, as the academic community comes to realize that input from target groups is crucial to the success of health interventions ^38^. Despite this fact, the bulk of the work in this area is still done with relatively little engagement, or a low level of participation of patients – who often have a minor advisory or consultation role, and who may be used in a tokenistic fashion ^39^. Engaging patients using a participatory approach is one way to meaningfully engage populations in all aspects of the research.

Research with community members who face significant social disadvantages can be challenging, as we demonstrated, with half of our participants facing acute health challenges that precluded their participation for a time. The social instability faced by this population can also hinder engagement in research, as was demonstrated by frequent changes in residence and telephone numbers during the study period. There is a multiplicity of other challenges with this kind of work, including difficulty in securing funding and getting approval from research ethics boards at the outset, when a discrete research question or methodology has not yet been identified. This required a flexible funder, and a very basic initial ethics application with numerous subsequent modifications as the study evolved. With respect to compensation, best practice in patient-oriented research states that participants should be compensated as they are providing time, knowledge and expertise ^40^, yet this needs to be balanced with the possibility that compensation may be coercive to a socially disadvantaged population, as it may provide an incentive for them to participate when in fact they do not wish to do so ^41^. Finally, this type of work is exceptionally time consuming for the investigators/group facilitators and requires the ability to manage group dynamics and individual personalities in a way that maintains the productivity of the group meetings.

Despite these challenges, there are a number of important benefits to using participatory approaches that researchers should consider. Firstly, because the community is involved with the planning and conducting of the research, one can be sure that the topics studied are of relevance to the communities affected and that researchers’ efforts will not simply serve their own academic interest, such as publications and presentations, but will additionally have meaningful practical value. Furthermore, by engaging community members early on, they will be empowered to help researchers communicate study findings and recommendations to decision makers.

Despite the many intrinsic strengths of this methodology, there are certainly limitations that warrant consideration. First, the sample size was small, even for a qualitative research study. Because of the dynamic and iterative nature of this work, we were unable to continue sampling or adding group members until saturation was reached. Therefore, it is unclear how our group members’ experiences relate to the broader population of patients living with diabetes and homelessness. While we had a diverse group, which included both men and women of multiple ethnic backgrounds, and a variety of diabetes-related presentations (i.e. complications, treatments, and diabetes duration), we certainly did not have representation from all groups. In particular, we did not capture the perspectives of those living with type 1 diabetes. Therefore, many potential concerns around insulin titration and hypoglycemia were not strongly represented in the knowledge that was co-created. Finally, because of the context-specific nature of this type of research, the generalizability of these findings to other settings is unclear.

This study offers valuable experience regarding the use of participatory patient-engagement research in studying diabetes experiences in a traditionally underserved population. Participatory research studies that engage similar populations should use methods that maximally involve patients as partners in the research process. This type of work is not without challenges, but has a number of advantages over traditional research approaches. We feel that this methodology can provide an important starting point for defining and understanding the priorities of communities with whom researchers can partner to improve healthcare service delivery. Researchers might consider starting research programs with community engaged work of this nature to help ensure that their study findings have maximal impact and result in interventions that meet the needs of communities.

## Data Availability

Data for this study are not publicly available.

## Acknowledgements

We would like to acknowledge the critical contributions of additional our group members who are not co-authors of this manuscript: Dwayne E. Hunte, Jasmine, Cat, Ozzy, and Georgina Bird. We thank them each for their willingness to participate and engage in the research process described. We thank the Interdisciplinary Chronic Disease Collaboration team: Corri Robb for transcription services, Sarah Gil for graphic design, and Patricia Wiebe for research administration. Linda Monteith-Gardner was instrumental in the photovoice project and Patricia O’Campo helped with the concept mapping. Nishan Zewge-Abubaker and Tadios Tibebu facilitated the evaluation focus group. We are grateful for the staff of the TD Centre for Learning for hosting this community-based research group.

## Data Availability

Data for this study are not publicly available.

## Funding

This work was supported by Alberta Innovates; the O’Brien Institute for Public Health Vulnerable Populations Research Fund; and the Cal Wenzel Family Cardiometabolic Fund for Research at the University of Calgary. The funders played no role in the data collection, analysis or interpretation.

## Authors’ relationships and activities

None of the authors have any conflicts of interest to declare.

## Contribution statement

The study was conceived by DC and RC with contributions from SW, GB and KM. The study topics were decided upon by AD, ML, MD in conjunction with DC and RC and the other members of the CDAC. Data collection was completed by DC, RC, AD, ML, MD. DC led the analysis of the data and wrote the first draft of the manuscript. All authors contributed significantly to data interpretation, critically revised and approved of the final submitted manuscript. DC is the guarantor for the manuscript.

## Notes

### Competing Interest Statement

The authors have declared no competing interest.

### Funding Statement

This work was supported by Alberta Innovates; the OBrien Institute for Public Health Vulnerable Populations Research Fund; and the Cal Wenzel Family Cardiometabolic Fund for Research at the University of Calgary. The funders played no role in the data collection, analysis or interpretation.

### Author Declarations

Ethics approval for this work was received from the Conjoint Health Research Ethics Board of the University of Calgary (REB# 18-1663), as well as the Research Ethics Board of St. Michaels Hospital, Unity Health Toronto (REB # 18-288).

